# The Real-World Clinical Benefits of GLP-1 Receptor Agonist Treatment

**DOI:** 10.1101/2025.10.24.25338256

**Authors:** Henriette Coetzer, Caroline Margiotta, Amy Marr, Razia Hashmi, Jason Kay, Vikrant Vats, Stuart Hagen, Stephanie Tomlin, Laura Anatale-Tardiff, David Wennberg

**Affiliations:** Blue Health Intelligence, Chicago, IL; Blue Cross Blue Shield Association, Chicago, IL

## Abstract

**Background:** The cardiovascular, metabolic and renal benefits of glucagon-like peptide-1 receptor agonists (GLP-1RAs) have been established in randomized trials with carefully selected patients. Real-world evidence of GLP-1RA effects on progression to end-stage renal disease (ESRD), end-stage liver disease (ESLD), and major adverse cardiovascular events (MACE) is scant.

**Methods:** We conducted a large retrospective cohort study of GLP1-RA use in adults using a national medical and pharmacy claims dataset. GLP-1RA users were stratified by diabetes status and propensity score-matched to controls. Outcomes were incident ESRD, ESLD, MACE overall and by treatment persistence (<1 year, 1–<2 years, ≥2 years). Cox models were used to estimate adjusted hazard ratios (HRs) and to calculate confidence intervals (CIs).

**Results:** In members with diabetes, GLP-1RA therapy was associated with reduced risks of ESRD (HR, 0.55; 95% CI, 0.49 to 0.61), ESLD (HR=0.66; 95% CI 0.60-0.73) and MACE (HR=0.90; 95% CI 0.86-0.93). In members without diabetes, while the incidence rates of adverse outcomes were lower, treatment with GLP-1RAs still resulted in lower risk of ESRD (HR=0.66, CI 0.46-0.94), ESLD (HR-0.76, CI 0.62-0.93) and MACE (HR=0.86, CI 92-0.99). Persistent GLP-1RA treatment (≥2 years) in members with diabetes resulted in the largest observed reduction in ESRD, ESLD, and MACE risk.

**Conclusions:** GLP-1RA treatment was associated with reduced incidence of kidney and liver failure and MACE. The impact of GLP-1RA treatment was greatest in members with diabetes and those persisting for ≥2 years.

## Introduction

Obesity and type-2 diabetes (T2DM) continue to drive rising morbidity, mortality, and healthcare costs in the United States.^1–6^ Glucagon-like peptide-1 receptor agonists (GLP-1RAs), originally developed for glycemic control in T2DM, have emerged as transformative therapeutics for multiple indications. ^7–11^ Randomized controlled trials (RCTs) including LEADER, SUSTAIN-6, and REWIND demonstrated GLP-1RA efficacy in reducing major adverse cardiovascular events (MACE), improving renal outcomes, and contributing to weight loss in patients with and without diabetes.^12^ These findings have rapidly expanded the clinical indications and utilization of GLP-1RA therapies.

Preclinical and translational studies find GLP-1RAs exert anti-inflammatory effects, enhance natriuresis and reduce oxidative stress, which improves endothelial function. These mechanisms appear to preferentially benefit microvasculature of the kidneys and brain, with less immediate impact on macrovascular territories such as coronary arteries.^13–17^ However, GLP-1RA clinical trials have consistently underrepresented older adults, individuals with advanced chronic kidney disease (CKD), and those with multiple chronic conditions - populations common in real world practice who may experience different risks, adherence challenges, and therapeutic benefits.

We evaluated the real-world association between GLP-1RA treatment and the incidence and timing of end-stage renal disease (ESRD), end-stage liver disease (ESLD), and MACE using a large national claims database in members with and without diabetes. Leveraging real-world data, we aim to extend the understanding of GLP-1RA therapeutic effects beyond the constraints of clinical trial populations—to inform clinical care, risk stratification and value-based risk sharing arrangements. Our primary hypothesis is that sustained GLP-1RA use reduces the cumulative incidence of ESRD, ESLD and MACE and delays their onset.

## Methods

### Ethics statement

This study was deemed exempt by the Sterling Institutional Review Board (protocol #12167) with waiver of informed consent and followed the Strengthening the Reporting of Observational Studies in Epidemiology (STROBE) reporting guidelines.

### Setting and Data Sources

We used the Blue Cross Blue Shield (BCBS) Association’s National Data Warehouse,^1^ containing medical claims data from participating BCBS plans. Adults aged 18-64 (January 1, 2016-August 31, 2024) residing in the United States with commercial medical and pharmacy insurance were potentially eligible (N=24,987,055; Table S1).

### Cohort Construction

Treated members had ≥1 pharmacy claims for a GLP-1RA product (tirzepatide, albiglutide, semaglutide, liraglutide, lixisenatide, dulaglutide, or exenatide) between January 1, 2017 and February 29, 2024, with ≥12 months baseline continuous medical and pharmacy enrollment and ≥6 months follow-up after the initial prescription fill (index date; N=1,725,097).

<u>Control members</u> had no prescription fills for any GLP-1RA products between January 1, 2017 and August 31, 2024 and ≥18 months of continuous medical and pharmacy enrollment (N=23,261,958). Controls were randomly assigned a proxy exposure or pseudo-index date within quarterly periods from January 1, 2017 through February 29, 2024.^18^

Members in both cohorts were classified as having a condition if they had ≥1 inpatient facility claims or ≥2 professional claims on separate days (excluding ambulance, DME, laboratory, and imaging claims) with relevant ICD-10 diagnoses in their baseline year. Both cohorts excluded members with GLP-1RA contraindications, high-cost conditions (e.g. cancers except squamous and basal cell carcinoma, mechanical ventilation > 96 hours), conditions limiting long-term treatment (e.g. dementia, end of life conditions), or bariatric procedures, yielding 936,777 treated members and 18,281,409 eligible controls (Table S1).

### Matching

Treated members were exact-matched to controls on gender, age group, index or pseudo-index year-quarter, and baseline diabetes and weight status. Propensity score matching (caliper=0.05 standard deviations of the logit) was then performed on baseline comorbidities (anxiety, depression, asthma, COPD, obstructive sleep apnea, hypertension, dyslipidemia, cerebrovascular disease, chronic kidney disease, coronary artery disease, heart failure (HF), early-stage liver disease, late-stage liver disease, alcohol or opiate use disorder, pancreatitis, psoriatic and rheumatic disease, and weight-bearing joint pain), race and ethnicity (RAND Bayesian Improved First Name Surname Geocoding imputation), and healthcare utilization (diabetes medications, cardiovascular medications, non-bariatric inpatient stays, and number of unique drug classes filled).^19^ A 1:1 match ratio without replacement yielded 742,824 matched pairs—413,404 with diabetes and 329,420 without diabetes. Additional information on the definition of baseline matching conditions appears in the Methodology Supplement.

### Clinical Outcomes

Clinical outcomes were identified using a combination of International Classification of Diseases, 10^th^ Revision diagnosis (ICD-10-CM) codes, Current Procedural Terminology (CPT) and Healthcare Common Procedure Coding System (HCPCS) codes, Diagnosis-Related Groups (DRGs), and revenue center codes.

Claims for diagnostic testing, imaging, durable medical equipment, or ambulance transport were excluded. The outcome onset date was defined as the earliest claim date meeting the outcome criteria within the 6-year follow-up period. Outcomes required ≥1 inpatient claim or ≥2 non-inpatient claims within 365 days except as noted for kidney dialysis and specified MACE components.

ESRD was identified by the earliest of kidney dialysis lasting ≥28 days or a kidney transplant.^20,21^ ESLD was identified by the earliest of hepatic failure, liver transplant, or hepatocellular carcinoma. MACE comprised: (1) ≥1 inpatient claim for HF, acute coronary syndrome (ACS) / unstable angina (UA), acute MI, stroke, or urgent/emergent coronary revascularization; or (2) ≥2 non-inpatient claims for acute MI or stroke.^22^

Outcomes were censored at the earliest of the end of follow-up, the first event, or 6 years post-index. To avoid misclassification of prevalent conditions, members with evidence of each outcome during the 12-month baseline or within 30 days after the index or pseudo-index date were excluded from analyses of that outcome.

### Follow-Up

Members were followed until disenrollment from medical or pharmacy benefits, the end of the study period (31 August 2024), or 6 years post-index or pseudo-index date. Median follow-up time was 20 months for treated members and 19 months for controls. Persistence of GLP-1RA treatment was assessed by aggregating all GLP-1RA treatment periods for each member, allowing gaps of up to 90 days; gaps exceeding 90 days indicated discontinuation. Persistence patterns differed by diabetes status: 52.4% of those with diabetes and 66.5% of those without diabetes persisted for <1 year (short-term users), 29.1% of those with diabetes and 27.5% of those without diabetes persisted for 1 to <2 years (moderate-term users), and 18.5% of those with diabetes and 6.0% of those without diabetes persisted for ≥2 years (long-term users; Table S3).

### Statistical Analysis

Analyses were conducted with R Studio Version 2024.04.2 using the matchit and survival packages. Baseline characteristics were summarized using means and 95% confidence intervals for continuous variables, and frequency distributions for categorical variables.

Cox proportional hazards models estimated hazard ratios (HRs) with 95% confidence intervals over the 6-year follow-up period, stratified by diabetes status. Kaplan-Meier survival models assessed time-to-event outcomes, with cumulative incidence (events per 100 person-months) estimates calculated as 1 minus the Kaplan-Meier survival estimator, assuming no competing risks. Log-rank tests compared survival differences between groups over the 6-year follow-up period. Proportional hazards assumptions for the Cox models were verified using scaled Schoenfeld residuals, where global and cohort-specific tests with non-significant p-values confirmed proportionality. Bonferroni adjustment addressed multiple comparisons for each outcome, where unadjusted p-values were compared to an adjusted threshold of 0.05 divided by the number of comparisons being made.

## Results

### Baseline characteristics

Matched treated and control members were well-balanced on demographic and clinical characteristics. In members with diabetes, the mean age was 52.4 years in the treated group and 52.5 years in the control group; 51.0% of each group were male (Table 1). Most had type 2 diabetes without complications (63.7%), and the most common comorbidities were hypertension (61.8% of treated vs 63.6% of controls) and dyslipidemia (72.0% of treated vs 73.4% of controls). Follow-up time varied by GLP-1RA treatment persistence. At least 3 years of follow-up were available for 34,188 short-term treated (<1 year), 11,280 medium-term treated (1 to <2 years), 41,284 long-term treated (≥2 years), and 72,805 matched controls (Table S4).

**Table 1:** Characteristics of Matched GLP-1RA Treated and Control Cohorts.

| Characteristic considered in the matching algorithm | Members with Diabetes |  |  | Members without Diabetes |  |  |
| --- | --- | --- | --- | --- | --- | --- |
|  | Treated | Controls | SMD | Treated | Controls | SMD |
|  | N = 413,404 | N = 413,404 |  | N = 329,420 | N = 329,420 |  |
| Age at Index (Years), mean (SD) | 52.4 (8.8) | 52.5 (8.9) | 0.01 | 46.7 (10.2) | 46.7 (10.3) | 0.01 |
| Gender (% Female) | 49.0% | 49.0% | 0 | 75.7% | 75.7% | 0 |
| **Race/Ethnicity** |  |  |  |  |  |  |
| White | 63.5% | 61.1% | 0.05 | 70.5% | 70.9% | 0.01 |
| Black | 8.3% | 8.8% | 0.02 | 7.2% | 7.2% | 0 |
| Hispanic | 10.7% | 11.7% | 0.03 | 7.1% | 7.1% | 0 |
| Other | 4.6% | 5.1% | 0.02 | 2.9% | 2.5% | 0.02 |
| Unknown | 12.8% | 13.3% | 0.01 | 12.3% | 12.3% | 0 |
| Type 1 Diabetes | 2.0% | 2.0% | 0 | 0.0% | 0.0% | 0 |
| Type 2 Diabetes with complications | 34.3% | 34.3% | 0 | 0.0% | 0.0% | 0 |
| Type 2 Diabetes without complications | 63.7% | 63.7% | 0 | 0.0% | 0.0% | 0 |
| Obesity | 35.7% | 35.7% | 0 | 45.1% | 45.1% | 0 |
| Liver Disease | 4.2% | 4.0% | 0.01 | 2.7% | 2.2% | 0.01 |
| Hypertension | 61.8% | 63.6% | 0.04 | 35.4% | 35.9% | 0.01 |
| Anxiety | 11.7% | 11.9% | 0.01 | 20.1% | 20.2% | 0.00 |
| Depression | 9.4% | 9.4% | 0.00 | 14.0% | 13.8% | 0.01 |
| Heart Failure | 2.1% | 2.1% | 0.00 | 0.6% | 0.3% | 0.03 |
| Coronary Artery Disease | 6.9% | 6.6% | 0.01 | 2.1% | 1.9% | 0.01 |
| Chronic Kidney Disease | 4.2% | 4.2% | 0 | 0.9% | 0.7% | 0.02 |
| Obstructive Sleep Apnea | 11.0% | 10.3% | 0.02 | 9.7% | 9.1% | 0.02 |
| **Prior Treatment** |  |  |  |  |  |  |
| Insulin | 13.6% | 12.9% | 0.02 | 0.0% | 0.0% | 0.00 |
| SGLT2 Inhibitors | 18.6% | 16.2% | 0.06 | 0.0% | 0.0% | 0.00 |
| Sulfonylureas<sup>a</sup> | 21.9% | 21.3% | 0.01 | 0.0% | 0.0% | 0.00 |
| DPP-4 Inhibitors | 10.5% | 9.9% | 0.02 | 0.0% | 0.0% | 0.00 |
| Biguanides | 72.4% | 73.8% | 0.03 | 2.3% | 1.3% | 0.08 |
| **Number of non-bariatric inpatient stays** |  |  |  |  |  |  |
| 0 | 95.0% | 95.2% | 0.01 | 97.6% | 97.8% | 0.01 |
| 1 | 4.2% | 3.9% | 0.01 | 2.0% | 1.9% | 0.01 |
| 2+ | 0.9% | 0.9% | 0 | 0.3% | 0.3% | 0.01 |
| **Number of other prescription classes used at baseline** |  |  |  |  |  |  |
| 0 | 5.2% | 4.7% | 0.02 | 4.5% | 4.6% | 0 |
| 1 to 3 | 30.4% | 29.8% | 0.01 | 25.0% | 24.9% | 0 |
| 4 to 6 | 29.3% | 29.7% | 0.01 | 28.0% | 28.1% | 0 |
| 7 to 9 | 17.9% | 18.3% | 0.01 | 20.1% | 20.2% | 0 |
| 10+ | 17.1% | 17.5% | 0.01 | 22.4% | 22.3% | 0 |

<sup>a</sup> The sulfonylureas category also includes meglitinide analogues and thiazolidinediones.

Treated and control members without diabetes had a mean age of 46.7 years, and 75.7% were female, 45.1% had a diagnosis of obesity or overweight.

Hypertension and dyslipidemia were the most common comorbidities, with similar prevalence between treated and control groups. At least 3 years of follow-up were available for 20,589 short-term treated, 5,252 moderate-term treated, 7,453 long-term treated, and 31,547 matched controls.

### End-Stage Renal Disease

Treated members with diabetes had a 45% lower risk of ESRD (p<0.001) compared to controls (Figure 1). The magnitude of the effect was similar for dialysis and kidney transplant, the two components of ESRD (Figure 1; Table 2). We found a strong ‘persistence response curve’ for ESRD in members with diabetes: those treated for ≥2 years had a 73% lower risk (p<0.001), members treated for 1-<2 years had a 55% lower risk (p<0.001) than controls, while members treated for <1 year had no significant risk reduction (Figure 2; Table 2). GLP-1RA treatment delayed the onset of ESRD: at 3 years, 0.42% of controls developed ESRD, an incidence rate delayed by 1.2 years in members with any GLP-1RA treatment and 2.7 years in members treated for ≥2 years.

**Fig 1.**
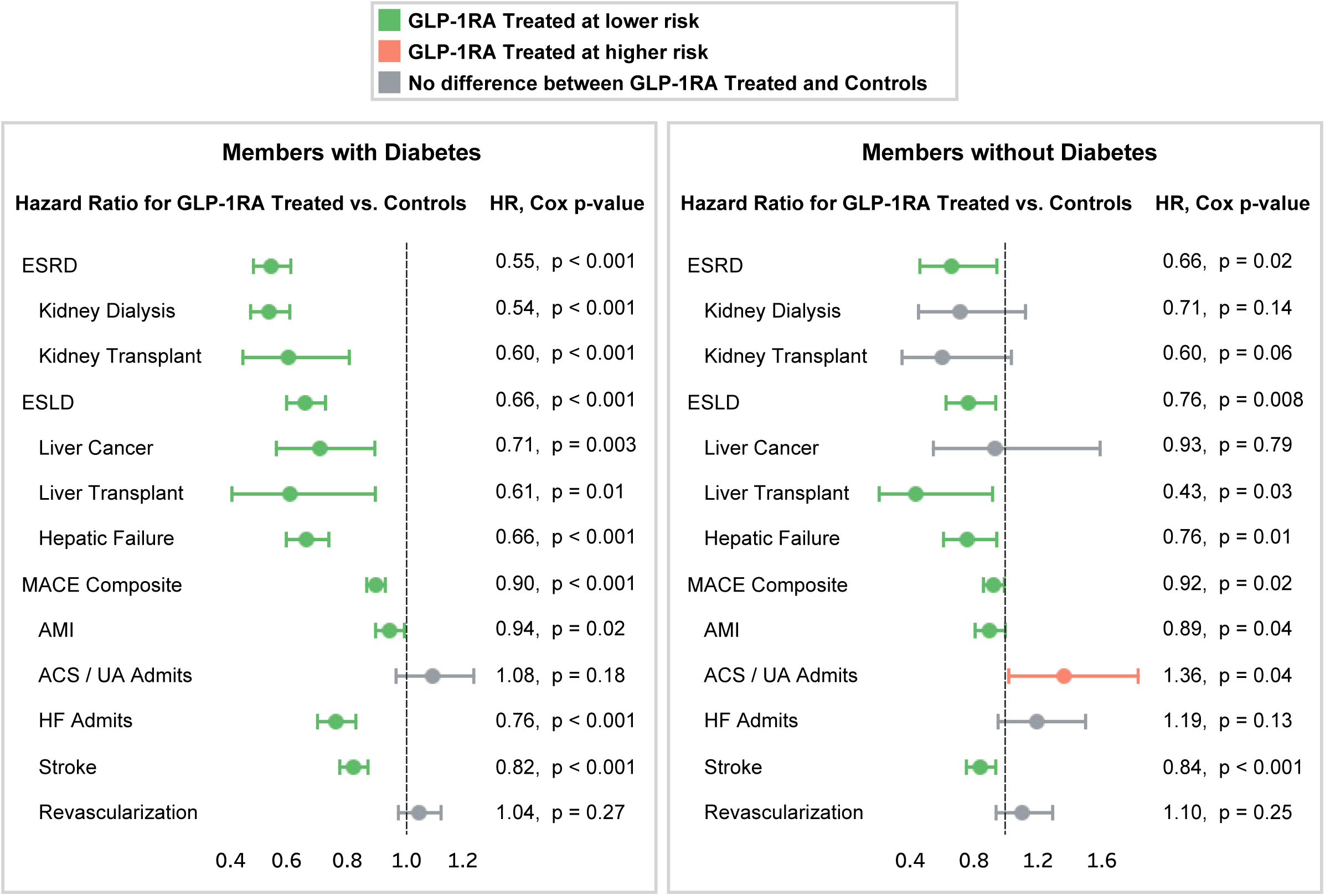
Hazard of Incident ESRD, ESLD, and MACE between GLP-1RA Treated and Controls, for Members with and without Diabetes. Hazard ratios (HR) and p-values were generated by Cox proportional hazards models over the 6-year member follow-up period. Statistical significance was established using a threshold of α = 0.05.

**Fig 2.**
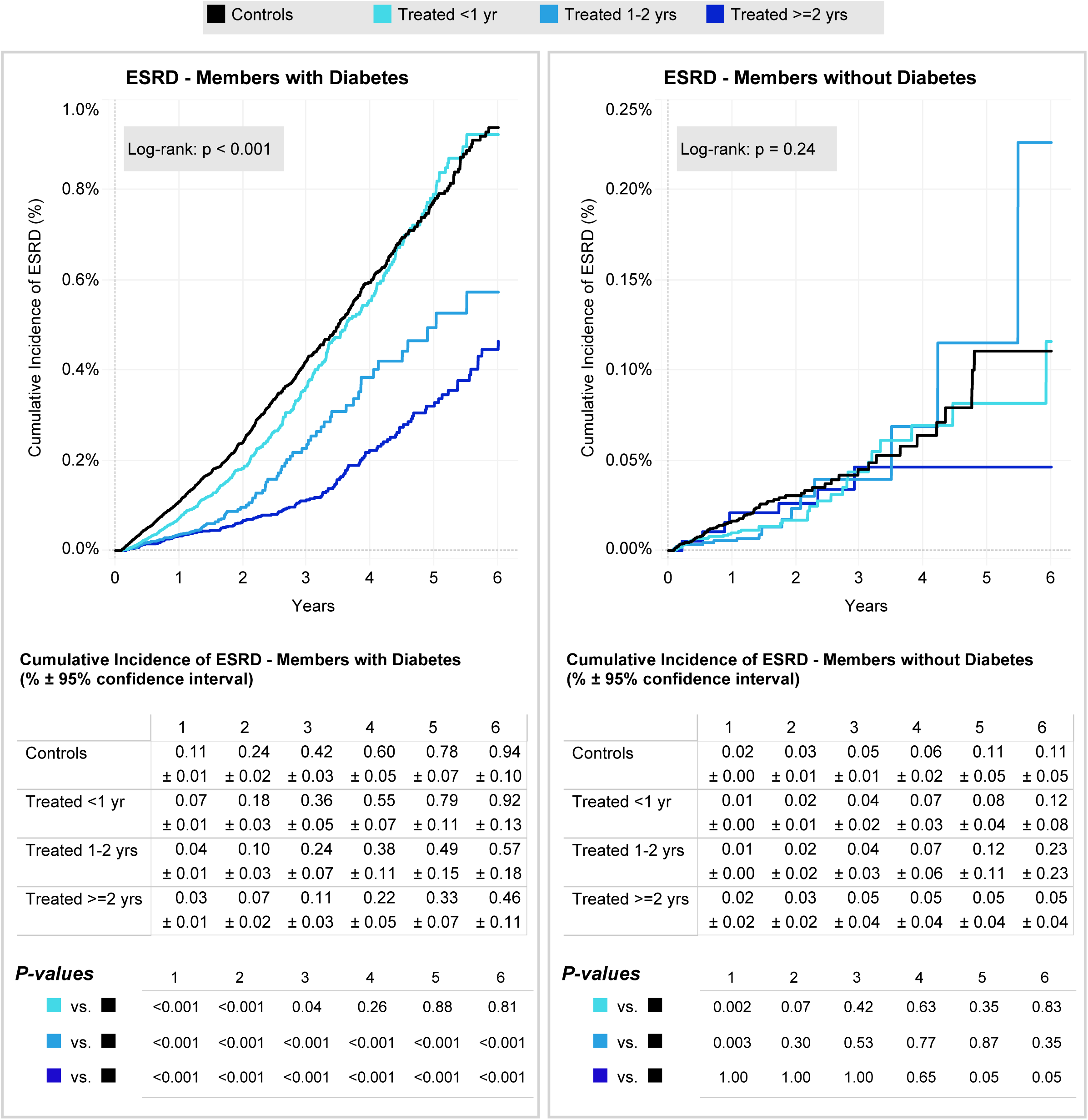
Cumulative Incidence of ESRD by 3 Levels of GLP-1RA Persistence and Untreated Controls, for Members with and without Diabetes. Log-rank p-value on the graphs shows that the cumulative incidence of ESRD varies by degree of persistence in members with diabetes but not significantly in members without diabetes. Tables below graphs show corresponding cumulative incidence rates (%) and 95% confidence bounds at one year timepoints. In the bottom table, raw p-values of Wald z-test comparisons between each GLP-1RA persistence stratum to the control group should be compared to a Bonferroni-adjusted threshold of α = 0.016 (0.05/3).

**Table 2:** 6-Year Cumulative Incidence and Hazard of End-Stage Renal Disease.

| Outcome | Degree of GLP-1 Persistence | Members with Diabetes |  |  |  |  | Members without Diabetes |  |  |  |  |
| --- | --- | --- | --- | --- | --- | --- | --- | --- | --- | --- | --- |
|  |  | Cumulative Incidence (% , ± 95% CI) | Cumulative Incidence P-Value | HR | 95% CI | Cox P-Value<sup>a</sup> | Cumulative Incidence (% , ± 95% CI) | Cumulative Incidence P-Value | HR | 95% CI | Cox P-Value<sup>a</sup> |
| End-Stage Renal Disease | Controls | 0.94±0.10 |  |  |  |  | 0.11±0.05 |  |  |  |  |
|  | Treated: Overall | **0.67±0.08** | **<0.001** | **0.55** | **0.49-0.61** | **<0.001** | 0.12±0.06 | 0.807 | 0.66 | 0.46-0.94 | 0.021 |
|  | Treated: <1 year | 0.92±0.13 | 0.809 | 0.82 | 0.70-0.96 | 0.012 | 0.12±0.08 | 0.831 | 0.62 | 0.4-0.96 | 0.031 |
|  | Treated: 1-<2 years | **0.57±0.18** | **<0.001** | **0.45** | **0.35-0.57** | **<0.001** | 0.23±0.24 | 0.345 | 0.82 | 0.41-1.67 | 0.591 |
|  | Treated: ≥ 2 years | **0.46±0.11** | **<0.001** | **0.27** | **0.21-0.35** | **<0.001** | 0.05±0.04 | 0.050 | 0.57 | 0.17-1.95 | 0.372 |
| Kidney Transplant | Controls | 0.12±0.03 |  |  |  |  | 0.03±0.02 |  |  |  |  |
|  | Treated: Overall | 0.11±0.03 | 0.660 | **0.60** | **0.45-0.81** | **0.001** | 0.03±0.02 | 1.000 | 0.60 | 0.35-1.03 | 0.064 |
|  | Treated: <1 year | 0.13±0.05 | 0.716 | 0.65 | 0.43-0.97 | 0.036 | 0.03±0.03 | 1.000 | 0.46 | 0.22-0.94 | 0.032 |
|  | Treated: 1-<2 years | 0.11±0.09 | 0.838 | 0.65 | 0.37-1.13 | 0.127 | 0.03±0.03 | 1.000 | 1.20 | 0.37-3.93 | 0.763 |
|  | Treated: ≥ 2 years | 0.09±0.06 | 0.366 | 0.47 | 0.25-0.88 | 0.019 | 0.03±0.02 | 1.000 | 0.67 | 0.19-2.36 | 0.530 |
| Dialysis | Controls | 0.89±0.10 |  |  |  |  | 0.08±0.04 |  |  |  |  |
|  | Treated: Overall | **0.63±0.07** | **<0.001** | **0.54** | **0.48-0.61** | **<0.001** | 0.10±0.06 | 0.606 | 0.71 | 0.45-1.12 | 0.140 |
|  | Treated: <1 year | 0.86±0.13 | 0.710 | 0.85 | 0.72-1.0 | 0.046 | 0.09±0.07 | 0.820 | 0.73 | 0.42-1.27 | 0.269 |
|  | Treated: 1-<2 years | 0.57±0.19 | 0.003 | **0.42** | **0.32-0.55** | **<0.001** | 0.21±0.24 | 0.304 | 0.77 | 0.34-1.75 | 0.533 |
|  | Treated: ≥ 2 years | **0.44±0.11** | **<0.001** | **0.25** | **0.19-0.33** | **<0.001** | 0.02±0.03 | 0.025 | - | - | - |

<sup>a</sup> Raw p-values for GLP-1 users overall were compared to a threshold of $\alpha=0.05$ . P-values by degree of GLP-1 persistence were compared to a Bonferroni-adjusted threshold of $\alpha=0.017$ (0.05/3).

In members without diabetes, there was a 34% reduction in ESRD risk (p=0.02) (Figure 1). Few members without diabetes had 6 years of follow-up, and cumulative incidence of ESRD was low among treated and controls (0.12% versus 0.11%; Table 2). We found no relationship between treatment persistence and risk of ESRD in members without diabetes (Figure 2; Table 2).

### End-Stage Liver Disease

GLP-1RA treatment was associated with lower ESLD risk in members with and without diabetes (Figure 1). For members with diabetes, any GLP-1RA treatment was associated with a 34% lower risk (p<0.001) of ESLD (Figure 1; Table 3). For members with diabetes, the magnitude of the effect was similar for all three components of the composite measure (Figure 1). In treated members without diabetes there was a 24% reduction in ESLD, although the absolute differences in ESLD incident rates were small, and only two of the component measures (liver transplant, hepatic failure) were significantly lower in treated members (Figure 1; Table 3).

**Table 3:** 6-Year Cumulative Incidence and Hazard of End-Stage Liver Disease.

| Outcome | Degree of GLP-1 Persistence | Members with Diabetes |  |  |  |  | Members without Diabetes |  |  |  |  |
| --- | --- | --- | --- | --- | --- | --- | --- | --- | --- | --- | --- |
|  |  | Cumulative Incidence (% , ± 95% CI) | Cumulative Incidence P-Value | HR | 95% CI | Cox P-Value<sup>a</sup> | Cumulative Incidence (% , ± 95% CI) | Cumulative Incidence P-Value | HR | 95% CI | Cox P-Value<sup>a</sup> |
| End-Stage Liver Disease | Controls | 1.08±0.11 |  |  |  |  | 0.26±0.07 |  |  |  |  |
|  | Treated: Overall | **0.86±0.09** | **0.002** | **0.66** | **0.60-0.73** | **<0.001** | 0.27±0.08 | 0.85 | 0.76 | 0.62-0.93 | 0.008 |
|  | Treated: <1 year | 0.98±0.13 | 0.257 | 0.96 | 0.84-1.11 | 0.594 | 0.30±0.11 | 0.54 | 0.95 | 0.74-1.22 | 0.705 |
|  | Treated: 1-<2 years | 1.01±0.28 | 0.649 | **0.57** | **0.47-0.70** | **<0.001** | 0.15±0.11 | 0.11 | **0.43** | **0.28-0.66** | **<0.001** |
|  | Treated: ≥ 2 years | **0.67±0.13** | **<0.001** | **0.37** | **0.30-0.45** | **<0.001** | 0.26±0.15 | 1.00 | 0.68 | 0.35-1.31 | 0.253 |
| Liver Cancer | Controls | 0.17±0.04 |  |  |  |  | 0.05±0.05 |  |  |  |  |
|  | Treated: Overall | 0.20±0.04 | 0.306 | 0.71 | 0.56-0.89 | 0.003 | 0.02±0.02 | 0.23 | 0.93 | 0.54-1.58 | 0.786 |
|  | Treated: <1 year | 0.24±0.08 | 0.103 | 1.17 | 0.84-1.64 | 0.348 | 0.03±0.03 | 0.45 | 1.64 | 0.85-3.19 | 0.143 |
|  | Treated: 1-<2 years | 0.23±0.13 | 0.384 | 0.65 | 0.41-1.03 | 0.069 | 0.00±0.00 | 0.03 | 0.25 | 0.05-1.18 | 0.080 |
|  | Treated: ≥ 2 years | 0.14±0.06 | 0.392 | **0.30** | **0.18-0.50** | **<0.001** | 0.01±0.02 | 0.11 | 0.17 | 0.02-1.38 | 0.097 |
| Liver Transplant | Controls | 0.09±0.04 |  |  |  |  | 0.02±0.03 |  |  |  |  |
|  | Treated: Overall | 0.06±0.02 | 0.176 | 0.61 | 0.41-0.89 | 0.011 | 0.02±0.01 | 1.00 | 0.43 | 0.21-0.91 | 0.028 |
|  | Treated: <1 year | 0.07±0.03 | 0.434 | 0.89 | 0.51-1.54 | 0.675 | 0.02±0.02 | 1.00 | 0.50 | 0.20-1.24 | 0.134 |
|  | Treated: 1-<2 years | 0.12±0.11 | 0.603 | 0.47 | 0.21-1.05 | 0.065 | 0.00±0.00 | 0.12 | 0.25 | 0.05-1.18 | 0.080 |
|  | Treated: ≥ 2 years | 0.05±0.03 | 0.089 | 0.39 | 0.18-0.85 | 0.017 | 0.02±0.03 | 1.00 | 1.00 | 0.06-15.99 | 1.000 |
| Hepatic Failure | Controls | 0.93±0.10 |  |  |  |  | 0.24±0.07 |  |  |  |  |
|  | Treated: Overall | 0.73±0.08 | **0.003** | **0.66** | **0.60-0.74** | **<0.001** | 0.25±0.08 | 0.85 | 0.76 | 0.61-0.94 | 0.012 |
|  | Treated: <1 year | 0.84±0.12 | 0.272 | 0.97 | 0.84-1.13 | 0.702 | 0.27±0.10 | 0.64 | 0.92 | 0.71-1.20 | 0.541 |
|  | Treated: 1-<2 years | 0.83±0.25 | 0.475 | **0.57** | **0.46-0.71** | **<0.001** | 0.14±0.11 | 0.14 | **0.42** | **0.27-0.68** | **<0.001** |
|  | Treated: ≥ 2 years | 0.59±0.12 | **<0.001** | **0.36** | **0.28-0.45** | **<0.001** | 0.25±0.15 | 0.90 | 0.81 | 0.39-1.69 | 0.578 |

<sup>a</sup> Raw p-values for GLP-1 users overall were compared to a threshold of $\alpha=0.05$ . P-values by degree of GLP-1 persistence were compared to a
Bonferroni-adjusted threshold of $\alpha=0.017$ (0.05/3).

The relationship between GLP-1RA persistence and reduced ESLD differed by diabetes status. For members with diabetes, a sustained effect was found only in members treated for ≥2 years (Figure 3; Table 3). For members without diabetes, we found a sustained reduction in ESLD only in those treated between 1-2 years (Figure 3; Table 3). GLP-1RA treatment delayed the onset of ESLD among members with diabetes: at 3 years, 0.48% of controls had developed ESLD, a threshold not reached by members with any GLP-1RA treatment until an additional 8 months had elapsed. The delay was significantly more pronounced with longer GLP-1RA treatment, with long-term treated individuals taking an additional 2.3 years to reach this threshold.

**Fig 3.**
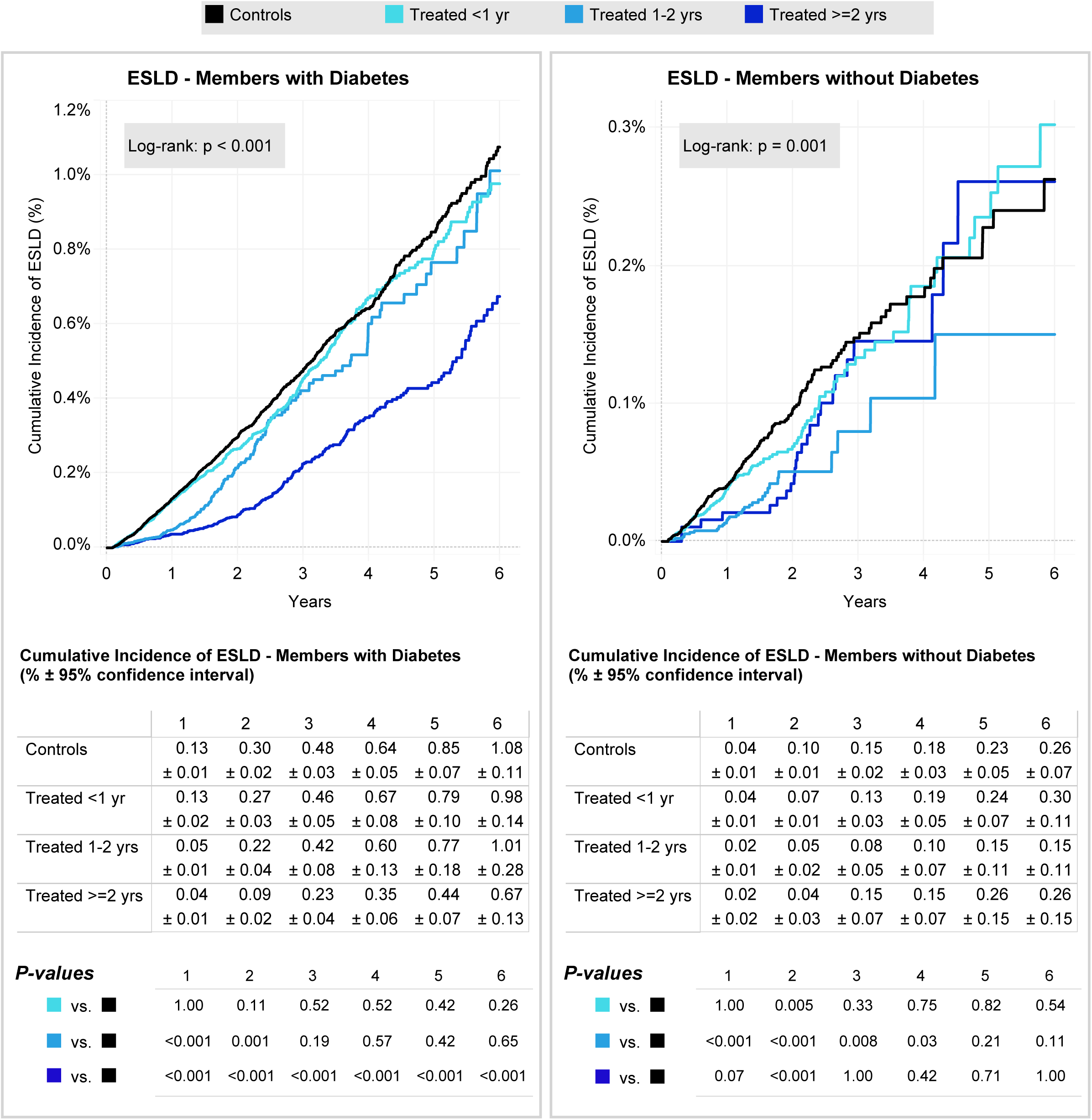
Cumulative Incidence of ESLD by 3 Levels of GLP-1RA Persistence and Untreated Controls, for Members with and without Diabetes. Log-rank p-value on the graphs shows that the cumulative incidence of ESLD varies by degree of persistence in members with and without diabetes. Tables below graphs show corresponding cumulative incidence rates (%) and 95% confidence bounds at one year time timepoints. In the bottom table, raw p-values of Wald z-test comparisons between each GLP-1RA persistence stratum to the control group should be compared to a Bonferroni-adjusted threshold of α = 0.016 (0.05/3).

### Major Adverse Cardiac Events

GLP-1RA treatment was associated with a lower risk of MACE in members with and without diabetes: relative risk was reduced by 10% and 8%, respectively (Figure 1). Among members with diabetes, risks of three of the five component events were reduced, the largest impact was on stroke and hospital admission for HF. Among members without diabetes, stroke risk was reduced by 16%, the largest component-specific effect (p<0.001).

MACE had the highest cumulative incidence of the three clinical outcome measures. By 6 years, 6.7% of untreated members with diabetes and 2.2% of members without diabetes had experienced a MACE (Figure 4; Table 4). Absolute risk differences favored GLP-1RA therapy; detailed estimates are provided in Table 4.

**Fig 4.**
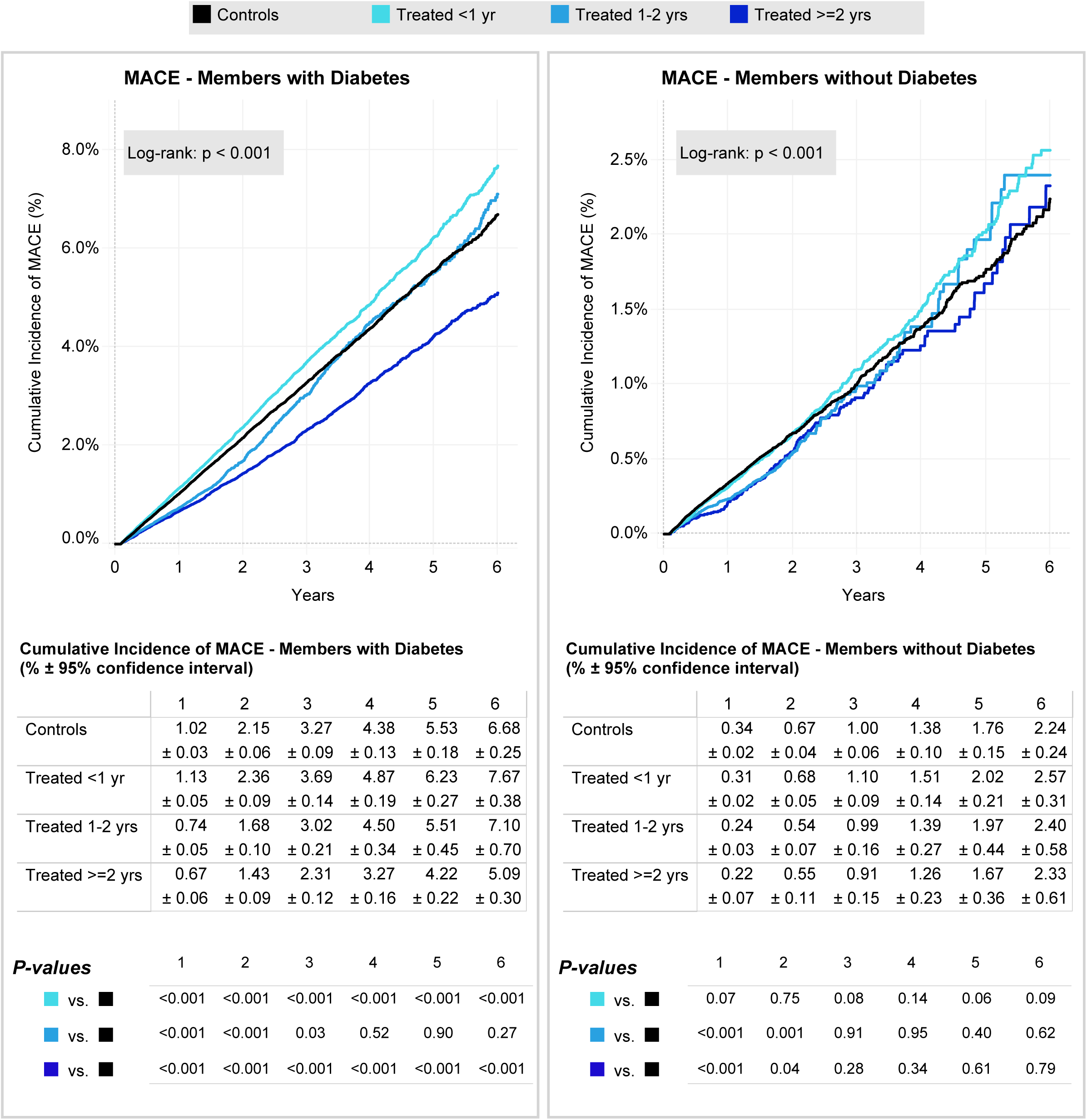
Cumulative Incidence of MACE by 3 Levels of GLP-1RA Persistence and Controls, for Members with and without Diabetes. Log-rank p-value on the graphs shows that the cumulative incidence of MACE varies by degree of persistence in members with and without diabetes. Tables directly below graphs show corresponding cumulative incidence rates (%) and 95% confidence bounds at one year time intervals. Raw p-values of Wald z-test comparisons between each GLP-1RA persistence stratum to the control group should be compared to a Bonferroni-adjusted threshold of α = 0.016 (0.05/3).

**Table 4:** 6-Year Cumulative Incidence and Hazard of Major Adverse Cardiac Events.

| Outcome | Degree of GLP-1 Persistence | Members with Diabetes |  |  |  |  | Members without Diabetes |  |  |  |  |
| --- | --- | --- | --- | --- | --- | --- | --- | --- | --- | --- | --- |
|  |  | Cumulative Incidence (% , ± 95% CI) | Cumulative Incidence P-Value | HR | 95% CI | Cox P-Value<sup>a</sup> | Cumulative Incidence (% , ± 95% CI) | Cumulative Incidence P-Value | HR | 95% CI | Cox P-Value<sup>a</sup> |
| MACE | Controls | 6.68±0.25 |  |  |  |  | 2.24±0.24 |  |  |  |  |
|  | Treated: Overall | 6.46±0.23 | 0.204 | **0.90** | **0.86-0.93** | **<0.001** | 2.49±0.25 | 0.152 | 0.92 | 0.86-0.99 | 0.019 |
|  | Treated: <1 year | **7.67±0.38** | **<0.001** | **1.17** | **1.12-1.23** | **<0.001** | 2.57±0.30 | 0.094 | 1.04 | 0.95-1.14 | 0.390 |
|  | Treated: 1-<2 years | 7.10±0.70 | 0.269 | **0.77** | **0.71-0.82** | **<0.001** | 2.40±0.58 | 0.615 | **0.72** | **0.62-0.82** | **<0.001** |
|  | Treated: ≥ 2 years | **5.09±0.30** | **<0.001** | **0.63** | **0.59-0.68** | **<0.001** | 2.33±0.61 | 0.788 | 0.81 | 0.65-1.02 | 0.070 |
| Acute MI | Controls | 3.30±0.19 |  |  |  |  | 1.10±0.17 |  |  |  |  |
|  | Treated: Overall | 3.14±0.16 | 0.203 | 0.94 | 0.89-0.99 | 0.020 | 1.14±0.16 | 0.741 | 0.89 | 0.80-0.99 | 0.037 |
|  | Treated: <1 year | **3.73±0.27** | **0.010** | **1.29** | **1.19-1.39** | **<0.001** | 1.23±0.21 | 0.352 | 0.96 | 0.84-1.09 | 0.503 |
|  | Treated: 1-<2 years | 3.69±0.55 | 0.190 | **0.77** | **0.70-0.86** | **<0.001** | 0.96±0.37 | 0.505 | 0.77 | 0.63-0.95 | 0.013 |
|  | Treated: ≥ 2 years | **2.43±0.20** | **<0.001** | **0.66** | **0.60-0.73** | **<0.001** | 0.96±0.33 | 0.459 | 0.86 | 0.61-1.21 | 0.385 |
| Acute Coronary Syndrome | Controls | 0.48±0.06 |  |  |  |  | 0.11±0.05 |  |  |  |  |
|  | Treated: Overall | **0.60±0.07** | **0.011** | 1.08 | 0.96-1.22 | 0.181 | **0.20±0.07** | **0.037** | 1.36 | 1.01-1.82 | 0.040 |
|  | Treated: <1 year | **0.71±0.12** | **0.001** | 1.31 | 1.11-1.56 | **0.002** | 0.16±0.07 | 0.255 | 1.38 | 0.95-2.01 | 0.090 |
|  | Treated: 1-<2 years | 0.59±0.21 | 0.326 | 0.85 | 0.67-1.09 | 0.196 | 0.18±0.14 | 0.349 | 1.18 | 0.67-2.09 | 0.564 |
|  | Treated: ≥ 2 years | 0.52±0.09 | 0.472 | 0.96 | 0.76-1.21 | 0.727 | 0.33±0.27 | 0.113 | 1.67 | 0.73-3.81 | 0.226 |
|  |  | Cumulative Incidence (% , ± 95% CI) | Cumulative Incidence P-Value | HR | 95% CI | Cox P-Value<sup>a</sup> | Cumulative Incidence (% , ± 95% CI) | Cumulative Incidence P-Value | HR | 95% CI | Cox P-Value<sup>a</sup> |
| Heart Failure | Controls | 1.46±0.13 |  |  |  |  | 0.24±0.09 |  |  |  |  |
|  | Treated: Overall | **1.15±0.10** | **<0.001** | **0.76** | **0.70-0.83** | **<0.001** | 0.28±0.08 | 0.518 | 1.19 | 0.95-1.49 | 0.134 |
|  | Treated: <1 year | 1.54±0.17 | 0.468 | 1.19 | 1.05-1.33 | 0.004 | 0.32±0.11 | 0.271 | **1.64** | **1.22-2.20** | **0.001** |
|  | Treated: 1-<2 years | 1.27±0.27 | 0.214 | **0.59** | **0.50-0.71** | **<0.001** | 0.27±0.21 | 0.800 | 0.82 | 0.51-1.31 | 0.407 |
|  | Treated: ≥ 2 years | **0.72±0.13** | **<0.001** | **0.37** | **0.30-0.44** | **<0.001** | 0.14±0.09 | 0.117 | 0.50 | 0.26-0.97 | 0.041 |
| Stroke | Controls | 2.48±0.15 |  |  |  |  | 0.91±0.15 |  |  |  |  |
|  | Treated: Overall | 2.43±0.15 | 0.645 | **0.82** | **0.77-0.87** | **<0.001** | 1.00±0.16 | 0.417 | 0.84 | 0.75-0.93 | **0.001** |
|  | Treated: <1 year | **2.92±0.25** | **0.004** | 1.04 | 0.96-1.13 | 0.303 | 1.01±0.19 | 0.419 | 0.99 | 0.86-1.14 | 0.889 |
|  | Treated: 1-<2 years | 2.52±0.43 | 0.863 | **0.74** | **0.66-0.83** | **<0.001** | 1.04±0.37 | 0.519 | **0.60** | **0.49-0.75** | **<0.001** |
|  | Treated: ≥ 2 years | **1.91±0.20** | **<0.001** | **0.58** | **0.51-0.65** | **<0.001** | 0.97±0.45 | 0.804 | 0.68 | 0.48-0.98 | 0.039 |
| Cardiac Revascularization | Controls | 1.71±0.13 |  |  |  |  | 0.43±0.10 |  |  |  |  |
|  | Treated: Overall | 1.86±0.12 | 0.104 | 1.04 | 0.97-1.11 | 0.272 | 0.51±0.11 | 0.292 | 1.10 | 0.94-1.29 | 0.254 |
|  | Treated: <1 year | **2.10±0.20** | **0.001** | **1.35** | **1.22-1.49** | **<0.001** | 0.50±0.12 | 0.393 | 1.14 | 0.94-1.40 | 0.186 |
|  | Treated: 1-<2 years | 2.19±0.41 | 0.030 | 0.92 | 0.80-1.06 | 0.250 | 0.47±0.26 | 0.780 | 1.00 | 0.73-1.37 | 1.000 |
|  | Treated: ≥ 2 years | 1.55±0.16 | 0.136 | **0.75** | **0.65-0.85** | **<0.001** | 0.63±0.31 | 0.234 | 1.06 | 0.65-1.74 | 0.803 |

<sup>a</sup> Raw p-values for GLP-1 users overall were compared to a threshold of $\alpha=0.05$ . P-values by degree of GLP-1 persistence were compared to a Bonferroni-adjusted threshold of $\alpha=0.017$ (0.05/3).

Among members with diabetes, treatment for ≥2 years was associated with a 1.5 percentage-point absolute reduction in 6-year MACE incidence–the largest absolute reduction observed across outcomes–and with delayed MACE onset. The controls’ 3-year cumulative MACE incidence of 3.3% was reached approximately 3 months later among those with any GLP-1RA exposure and 1.0 year later among those treated for ≥2 years. Among members without diabetes, MACE rates did not differ significantly after the 2^nd^ year of follow-up regardless of treatment persistence.

## Discussion

Our study is one of the largest published on the clinical impact of GLP-1RA use in actual practice. For members with diabetes, we find treatment with GLP-1RAs substantially decreases the risk of developing ESRD, ESLD or experiencing a MACE event. Members with diabetes are at much greater risk of developing ESRD, ESLD or experiencing a MACE event at baseline and this is reflected in the higher absolute reduction of these outcomes compared to members without diabetes. For members without diabetes, while the effects are more modest, we still find that treated members have lower rates of all three major outcomes. Our other major finding is that members with longer persistence are more likely to benefit. These results extend randomized-trial findings to unselected, working-age adults and quantify—for the first time in routine practice—the multi-year deferral of kidney, liver, and composite cardiovascular events.

Cardiovascular outcomes trials (including LEADER, SUSTAIN-6, and REWIND) have demonstrated that GLP-1RAs reduce the incidence of MACE and slow the progression of CKD. However, these trials enrolled highly selected patient populations and did not measure hard ESLD end points or time-to-event delays.^11–13^ The 45% relative reduction in ESRD observed in our study aligns with the 45% decline in the renal composite reported for dulaglutide in REWIND, our accompanying finding of a 2.7-year delay in ESRD represents a new clinically meaningful finding. At the population level, treating 250 treatment-adherent patients for a year prevents one renal-failure case—an effectiveness comparable to statins for secondary prevention of MI.

Real-world data on ESLD have been sparse and heterogeneous; the present study demonstrates a 34% reduction in hazard and a 2.3-year delay in progression to hepatic failure or transplant.^23^ The attenuation of acute MI risk—non-significant overall but reduced by 34% with long-term GLP-1RA exposure—mirrors trial-based observations that coronary benefit may accrue more slowly than cerebrovascular or heart-failure benefit, potentially reflecting differing pathophysiologic mechanisms and persistence of exposure requirements.

Longer follow-up studies are recommended to evaluate the impact of GLP- 1RAs on people without diabetes. In this study, the follow-up time and smaller absolute event rates in members without diabetes yielded wide confidence intervals around HR estimates. For approximately every 235 patients with diabetes who stay on a GLP-1RA for ≥12 months, one case of end-stage renal disease is averted within 6 years, and for liver failure the figure is approximately 270. Combined with the 2.7 year delay in dialysis and the 2.3 year delay in transplant-eligible ESLD, GLP1-RA treatment could result in significant reductions in medical costs for these rare but serious outcomes.^24,25^

This study has several limitations. Semaglutide was approved for diabetes in 2017 and in 2021 for obesity. Consequently, the number of treated individuals without diabetes available for analysis was lower than those with diabetes. However, this is still one of the largest studies to date of the impact of GLP-1RAs on people without diabetes. While diabetes is consistently coded in claims data, obesity is not.^26^ Our decision to consider all members without diabetes as a single cohort should be evaluated through comparing clinical data, such as electronic medical records, with claims data. Finally, the study was limited to members receiving GLP-1RAs via insurance benefits. Untreated members could have accessed GLP-1RAs via alternate mechanisms such as compounding pharmacies. Any misclassification would attenuate the observed differences between treated and untreated members.

We found that in members with diabetes persistent GLP-1RA therapy was associated with a reduced incidence and delayed onset of kidney and liver failure, as well as reduced rates of stroke and HF hospitalization. These findings provide real-world evidence of both clinical benefit and potential economic value among select patient population within a commercially insured cohort. Our study supports guideline recommendations to prioritize GLP-1RAs for high-risk patients and underscore the importance of adherence programs capable of supporting patients to persist with therapy for ≥1 year.

## Data Availability

Individual participant data will not be made available, in compliance with the data use agreement with the Blue Cross Blue Shield Association.

## Author Contributions

AM and CM had full access to all of the data in the study and take responsibility for the integrity of the data and the accuracy of the data analysis.

## Concept and design

HC, DW, AM, CM

## Acquisition, analysis, or interpretation of data

HC, AM, CM, DW

## Drafting of the manuscript

HC, AM, CM, LAT, ST

## Critical review of the manuscript for important intellectual content

HC, RH, JK, VV, SH, LAT, ST

## Statistical analysis

AM, CM

## Obtained funding

HC

## Administrative, technical, or material support

RH, JK, VV, SH, LAT

## Supervision

HC, DW

## AI Used in Manuscript Preparation

The drafting and refinement of this manuscript were supported by OpenAI’s ChatGPT, which assisted in enhancing the language and clarity of the text.

## Disclaimer

The content is solely the responsibility of the authors and does not necessarily represent the official views of the funding entity.

## Data Sharing Statement

Data will not be shared.

## Additional Contributions

The authors acknowledge Bob Darin (concept, design, review), Bolatito Adepoju (editing) and Thomas Parakilatu, Neha Jajodia, and Jen Zhong (data management) for their valuable contributions to this project.

## Footnotes

1 The National Data Warehouse (NDW) is a national data asset managed by BCBSA that houses BCBS Plan submitted data (e.g., medical claims, membership data) to support healthcare operations, program capabilities, and analytics.

