## Supplementary Tables for "The Real-World Clinical Benefits of GLP-1 Receptor Agonist Treatment" (from another version of this preprint: version 1, 2025-10-25)

**Authors:** Henriette Coetzer, MD[1], Caroline Margiotta, MA[1], Amy Marr, PhD[1], Razia Hashmi, MD[2], Jason Kay, PharmD[2], Vikrant Vats, PhD[2], Stuart Hagen, PhD[2], Stephanie Tomlin, MS[1], Laura Anatale-Tardiff, MPH[1], David Wennberg, MD[1]

**Affiliations:** 1) Blue Health Intelligence, Chicago, IL; 2) Blue Cross Blue Shield Association, Chicago, IL

| **Table of Contents** |
|---|
| Table S1a: GLP-1RA treated cohort identification and retention |
| Table S1b: Control cohort identification and retention |
| Table S2: Pre-matching characteristics of GLP-1RA treated and control cohorts |
| Table S3: GLP-1RA Persistence by Diabetes Status |
| Table S4: Follow-Up Time by GLP-1RA Persistence and Diabetes Status |

**Table S1a: GLP-1RA treated cohort identification and retention**

| Criteria | Members remaining | |
|---|---|---|
| | N | % of starting population |
| Members aged 18-64 years | 1,725,097 | 100.0% |
| Minimum of 12 months of continuous enrollment in primary medical and pharmacy coverage prior to index date and 6 months of continuous enrollment in primary medical and pharmacy coverage on and following index date | 1,018,206 | 59.0% |
| Members remaining after exclusions | 936,777 | 54.3% |

| Table S1b: Control cohort identification and retention | | |
|---|---|---|
| Criteria | Members remaining | |
| | N | % of starting population |
| Members aged 18-64 years of age | 23,261,958 | 100.0% |
| Minimum of 12 months of continuous enrollment in primary medical and pharmacy coverage prior to pseudo-index date and 6 months of continuous enrollment in primary medical and pharmacy coverage on and following pseudo-index date | 22,716,175 | 97.7% |
| Members remaining after exclusions | 18,281,409 | 78.6% |

**Table S2: Pre-matching characteristics of GLP-1RA treated and control cohorts**

| Characteristic considered in the matching algorithm | Full Population | | |
|---|---|---|---|
| | Treated | Controls | SMD |
| **Age at Index** | 32.8 | 24.8 | 0.67 |
| **Gender** | 60.4% | 52.1% | 0.17 |
| **Race/Ethnicity** | | | |
| White | 66.0% | 68.3% | 0.15 |
| Black | 8.1% | 5.5% | |
| Hispanic | 9.3% | 7.6% | |
| Other | 3.7% | 5.8% | |
| Unknown | 12.9% | 12.8% | |
| Type 1 Diabetes | 1.6% | 0.5% | 0.11 |
| Type 2 Diabetes with complications | 27.6% | 1.2% | 0.81 |
| Type 2 Diabetes without complications | 35.3% | 3.0% | 0.90 |
| Obesity | 44.7% | 9.1% | 0.89 |
| Liver Disease | 0.8% | 0.4% | 0.06 |
| Hypertension | 54.8% | 15.5% | 0.90 |
| Anxiety | 15.8% | 10.0% | 0.17 |
| Depression | 12.2% | 6.4% | 0.20 |
| Heart Failure | 1.5% | 0.3% | 0.13 |
| Coronary Artery Disease | 5.3% | 1.2% | 0.23 |
| Chronic Kidney Disease | 3.2% | 0.5% | 0.20 |
| Obstructive Sleep Apnea | 11.9% | 2.3% | 0.38 |
| Prior Treatment | | | |
| Insulin | 12.0% | 0.8% | 0.47 |
| SGLT2 Inhibitors | 14.8% | 0.5% | 0.56 |
| Sulfonylureas[a] | 17.0% | 0.7% | 0.60 |
| DPP-4 Inhibitors | 9.6% | 0.3% | 0.44 |
| Biguanides | 48.2% | 3.0% | 1.21 |
| Number of Non-Bariatric Inpatient Stays | | | |
| 0 | 95.6% | 98.1% | 0.14 |
| 1 | 3.6% | 1.6% | |
| 2+ | 0.7% | 0.3% | |
| Number of other prescription classes used at baseline | | | |
| 0 | 4.4% | 18.4% | 0.77 |
| 1 to 3 | 25.9% | 43.3% | |
| 4 to 6 | 28.3% | 22.5% | |
| 7 to 9 | 19.6% | 9.5% | |
| 10+ | 21.8% | 6.3% | |

[a] The sulfonylureas category also includes meglitinide analogues and thiazolidinediones.

**Table S3: GLP-1RA Persistence by Diabetes Status**

| GLP-1RA Persistence Cohort | Members with Diabetes | | | | Members without Diabetes | | | |
|---|---|---|---|---|---|---|---|---|
| | GLP-1RA Treated N = 413,404 | | Controls N = 413,404 | | GLP-1RA Treated N = 329,420 | | Controls N = 329,420 | |
| | N | % | N | % | N | % | N | % |
| Treated: <1 year | 216,729 | 52.4% | | | 219,036 | 66.5% | | |
| Treated: 1-<2 years | 120,198 | 29.1% | | | 90,744 | 27.5% | | |
| Treated: >= 2 years | 76,477 | 18.5% | | | 19,640 | 6.0% | | |
| Controls | | | 413,404 | 100.0% | | | 329,420 | 100.0% |

**Table S4: Follow-Up Time by GLP-1RA Persistence and Diabetes Status**

| Members remaining at the end of each year | Members with Diabetes | | | | | | | | Members without Diabetes | | | | | | | |
|---|---|---|---|---|---|---|---|---|---|---|---|---|---|---|---|---|
| | Treated: <1 year | | Treated: 1-<2 years | | Treated: >= 2 years | | Controls | | Treated: <1 year | | Treated: 1-<2 years | | Treated: >= 2 years | | Controls | |
| | N | % of starting population | N | % of starting population | N | % of starting population | N | % of starting population | N | % of starting population | N | % of starting population | N | % of starting population | N | % of starting population |
| Index | 220,535 | - | 119,275 | - | 73,594 | - | 413,404 | - | 220,361 | - | 90,008 | - | 19,051 | - | 329,420 | - |
| 1 Year | 125,184 | 56.8% | 119,275 | 100.0% | 73,594 | 100.0% | 302,972 | 73.3% | 148,789 | 67.5% | 90,008 | 100.0% | 19,051 | 100.0% | 244,599 | 74.3% |
| 2 Years | 63,327 | 28.7% | 21,840 | 18.3% | 73,594 | 100.0% | 141,953 | 34.3% | 49,230 | 22.3% | 16,528 | 18.4% | 19,051 | 100.0% | 80,141 | 24.3% |
| 3 Years | 34,188 | 15.5% | 11,280 | 9.5% | 41,284 | 56.1% | 72,805 | 17.6% | 20,589 | 9.3% | 5,252 | 5.8% | 7,453 | 39.1% | 31,547 | 9.6% |
| 4 Years | 19,418 | 8.8% | 5,958 | 5.0% | 23,124 | 31.4% | 38,966 | 9.4% | 10,880 | 4.9% | 2,511 | 2.8% | 3,339 | 17.5% | 15,974 | 4.8% |
| 5 Years | 10,960 | 5.0% | 3,266 | 2.7% | 12,583 | 17.1% | 20,669 | 5.0% | 5,930 | 2.7% | 1,340 | 1.5% | 1,636 | 8.6% | 8,590 | 2.6% |
| 6 Years | 5,404 | 2.5% | 1,521 | 1.3% | 5,478 | 7.4% | 9,348 | 2.3% | 2,907 | 1.3% | 609 | 0.7% | 704 | 3.7% | 4,120 | 1.3% |
